# Towards biomarkers for outcomes after pancreatic ductal adenocarcinoma and ischemic stroke, with focus on (co-)morbidity and aging / cellular senescence (SASKit): protocol for a prospective cohort study

**DOI:** 10.1101/2020.04.09.20037010

**Authors:** Larissa Henze, Uwe Walter, Hugo Murua Escobar, Christian Junghanß, Robert Jaster, Rüdiger Köhling, Falko Lange, Ali Salehzadeh-Yazdi, Olaf Wolkenhauer, Mohamed Hamed, Israel Barrantes, Daniel Palmer, Steffen Möller, Axel Kowald, Nicole Heussen, Georg Fuellen

**Affiliations:** Rostock University Medical Center, Department of Hematology, Oncology and Palliative Medicine, Department of Medicine III, Rostock, Germany; Rostock University Medical Center, Department of Neurology, Rostock, Germany; Rostock University Medical Center, Department of Gastroenterology, Rostock, Germany; Rostock University Medical Center, Oscar Langendorff Institute of Physiology, Rostock, Germany; University of Rostock, Department of Systems Biology and Bioinformatics, Rostock, Germany; Rostock University Medical Center, Institute for Biostatistics and Informatics in Medicine and Ageing Research, Rostock, Germany; RWTH Aachen, Department of Medical Statistics, Aachen, Germany

## Abstract

Aging-related processes such as cellular senescence are believed to underlie the accumulation of diseases in time, causing (co-)morbidity, including cancer, thromboembolism and stroke. Intervening into these processes may delay, stop or reverse morbidity. To study the link between (co-)morbidity and aging, by exploring biomarkers and molecular mechanisms of disease-triggered deterioration, we will recruit 50 patients with pancreatic ductal adenocarcinoma, 50 patients with (thromboembolic) ischemic stroke and 50 controls, at Rostock University Medical Center. We will gather routine blood data, clinical performance measurements and patient-reported outcomes at up to 9 points in time, and in-depth transcriptomics & proteomics at two early time points. Aiming for clinically relevant biomarkers, the primary outcome is a composite of probable sarcopenia, clinical performance (described by ECOG Performance Status for patients with pancreatic ductal adenocarcinoma and the Modified Rankin Scale for patients with stroke) and quality of life. Further outcomes cover other aspects of morbidity such as cognitive decline, and of comorbidity such as vascular or cancerous events. The data analysis is comprehensive in that it includes biostatistics & machine learning, both following standard role models & additional explorative approaches. *Predictive* biomarkers for interventions addressing senescence may become available if the biomarkers that we find are predominantly related to aging / cellular senescence. Similarly, *diagnostic* biomarkers will be explored for their relationship to aging / cellular senescence. Our findings will require validation in independent studies, and our dataset shall be useful to validate the findings of other studies. In some of the explorative analyses, we shall include insights from systems biology modeling as well as insights from preclinical animal models. We humbly suggest that our detailed study protocol and data analysis plan may also guide other biomarker exploration trials.

**In Brief:** The SASKit (“Senescence-Associated Systems diagnostics Kit for cancer and stroke”) study primarily aims to discover novel biomarkers for deterioration of health and (co-)morbidities triggered by pancreatic ductal adenocarcinoma or ischemic stroke.

## Introduction

### Study Rationale and Aims

The primary aim of the SASKit (“Senescence-Associated Systems diagnostics Kit for cancer and stroke”) study is to discover a set of molecular biomarkers for outcomes after pancreatic ductal adenocarcinoma (PDAC) and ischemic stroke (IS), which are specifically useful to predict disease-triggered deterioration of health (“disease deterioration” for short) in terms of probable sarcopenia ^1^, reduced clinical performance and quality of life (QoL). The outcomes also include the (co-)morbidity of vascular events (here defined as stroke, myocardial infarction, and venous or arterial thromboembolism) in patients with PDAC, which are observed frequently apart from sarcopenia. Also included is the (co-)morbidity of any kind of cancer and of cognitive decline following IS. Moreover, we consider mortality, as the most canonical outcome. Following up on the primary aim, we will investigate the nature of the molecular biomarkers to find out whether cellular senescence and other aging-associated processes are contributing to disease deterioration. As a secondary aim, we will search for *diagnostic* biomarkers related to cellular senescence and other aging-related processes that may differentiate healthy controls from PDAC or IS patients. Therefore, in the following we motivate our study by describing the prevalence and the outcomes of PDAC and IS, the known predictors of these outcomes, and the specific prevalence of co-morbidity and known predictors for this co-morbidity. The role of cellular senescence in aging and disease is described in Box 1. The background of the cancerous and vascular comorbidity is described in Box 2. Avoiding unclear or circular terminology, we define a biomarker in a very general fashion, simply as a feature (data point) f_1_ that successfully predicts another feature f_2_ at a later time-point ^2^, in a biomedical context. Here, features may be composite ones, based on the measurement of individual features. Often, feature f_1_ refers to molecular data, while feature f_2_ refers to phenotypic data, such as clinical outcomes. Ultimately, we aim to identify biomarkers that are easy to measure, and that are then validated in other studies to predict a clinically relevant outcome.

#### Box 1 Aging and cellular senescence

Extra lifetime gained over the last century led to the widespread emergence of age-related diseases that are rarely seen in younger people. Older patients are thus more likely to display several comorbidities, which makes treatment difficult and expensive. Over the last years, strong evidence has accumulated that the presence of senescent cells (i.e. non-dividing, arrested but metabolically active cells that escape apoptosis) is causally involved in diseases such as atherosclerosis, cancer, fibrosis, pancreatitis, osteoarthritis, Alzheimer disease and metabolic disorders ^43 44^. Evidence that senescent cells are not only correlated with aging and diseases, but are instead causally involved, comes from recent studies, which transplanted senescent cells from old into young mice ^45^. This resulted in persistent functional impairment as well as spread of cellular senescence to host tissues. Another strong line of evidence comes from experiments that actually removed senescent cells from aged mice by *senolytics* ^45-47^. In each case an increase in lifespan and a delay of typical age related diseases was observed. Most recently, the results of human pilot trials of putative senolytic treatments in case of idiopathic pulmonary fibrosis and osteoarthritis have been reported. One team ^48^ treated idiopathic pulmonary fibrosis patients with dasatinib and quercetin and demonstrated safety as well as notable improvements in some physical abilities. Furthermore, a human phase-1 study demonstrated that a senolytic compound, which was applied locally in patients with osteoarthritis of the knee, was safe and well-tolerated ^49^. A clinically meaningful improvement in several measures, including pain, function, as well as modulation of certain senescence-associated secretory phenotype (SASP) factors and disease-related biomarkers was observed after a single dose.

### Pancreatic ductal adenocarcinoma: prevalence and outcomes

The incidence of pancreatic cancer is increasing; in 2017 the global incidence was 5.7 per 100,000 person-years ^3^. Age is the most important risk factor, and incidence peaks at 65 to 69 years in males and 75 to 79 years in females ^3^. Pancreatic ductal adenocarcinoma (PDAC) is the most common histological type of pancreatic cancer^4^. The disease is characterized by late clinical presentation ^5^, early metastases and poor prognosis, with a one-year survival rate in Europe of only 15% ^6^. Many patients have unresectable disease at the time of diagnosis, either as locally advanced disease or already with metastases. Therefore therapy is palliative consisting of chemotherapy and/or best supportive care. Disease deterioration with weight loss and low muscle strength, that is, cachexia and sarcopenia ^7^, will follow, for some patients rapidly (within a few weeks) and for others during a longer interval of one or two years. Recent developments in oncology have not shown much benefit in clinical trials of patients with PDAC ^8^. Inflammation, desmoplasia and early metastases are deemed responsible for the difficulties in targeting the disease. Moreover, vascular events are frequent problems in the course of PDAC and may contribute to disease deterioration or early death. Venous thromboembolism is the most common event occurring in up to 34% of patients with metastatic PDAC ^9 10^, but arterial ischemic events, like stroke, are also reported ^11-14^, see also Box 2. Therefore, deterioration and mortality in PDAC can not only be explained by tumor progression as such, but other factors like sarcopenia/cachexia and vascular events contribute as well. Furthermore, we suggest that the underlying cause of all these factors are aging-related processes such as cellular senescence and chronic inflammation.

#### Box 2 Cellular senescence and the comorbidity of cancer and vascular events

Some cancers such as PDAC can trigger vascular events by hyper-coagulation, reflecting Trousseau’s syndrome first reported 150 years ago ^11^. In turn, strong associations between coagulation, cellular senescence and the SASP were demonstrated recently ^50^. While cellular senescence can suppress PDAC and cancerous proliferation in general, it also triggers tumor progression by fostering inflammatory processes, including the SASP, while on the other hand, after ischemic stroke, it attenuates recovery^51-55^. For both diseases, causal influences can be traced back to molecular determinants: PAI-1 (also known as SERPINE1 and part of the SASP) is involved in cancer-triggered thromboembolism ^52 54^ and stroke recovery in animals ^56^. Other proteins involved in cellular senescence, specifically inflammatory cytokines such as IL6, and the lesser known osteopontin and gelsolin, are also markers for both PDAC and stroke ^57-60^. The cyclin-dependent kinase CDK5 ^61^ is implicated in the progression of PDAC as well as in the recovery from stroke ^55 62^. Moreover, apart from being genetic risk factors ^63 64^, the most prominent drivers of cellular senescence (p16/CDKN2A and p21/CDKN1A) also promote PDAC progression ^65^ and endothelial embolic and arteriosclerotic mechanisms of stroke ^66^. Finally, two small-molecule interventions into cellular senescence, fisetin and quercetin, are both potential treatments of both PDAC and stroke. In case of stroke, the blood-brain-barrier is passed by quercetin which improves stroke outcome ^67^. In case of PDAC it was observed that quercetin inhibits pancreatic cancer growth *in-vitro* and *in-vivo* ^68^. Fisetin is found in various fruits (especially strawberries) and it is chemically similar to quercetin, with strong putative senolytic effects, extending lifespan of mice even when intervention with fisetin started only at an advanced age ^69^. In a study involving nude mice implanted with prostate cancer cells, treatment with fisetin significantly retarded tumor growth ^70^. Also, in case of lung cancer, there is evidence for the beneficial effects of fisetin. One study showed that fisetin provides protection against benzo(a)pyrene [B(a)P]-induced lung carcinogenesis in albino mice ^71^ and another *in vivo* study demonstrated the synergistic effects of fisetin and cyclophosphamide in reducing the growth of lung carcinoma in mice ^72^. Several other studies have also demonstrated its anticarcinogenic, neurotrophic and anti-inflammatory effects that are beneficial in numerous diseases, including pancreatic cancer and stroke ^73^.

### Pancreatic ductal adenocarcinoma: known biomarkers and clinical scores

In PDAC patients there is a lack of established scores describing the risk of disease deterioration and the risk of sarcopenia/cachexia in particular. Referring to the endpoint of overall survival, some recent studies tried to establish inflammation-based scores to better characterize outcome in PDAC. In a retrospective analysis of 386 patients with PDAC of different stages, CRP/Alb ratio, neutrophil– lymphocyte ratio (NLR), platelet–lymphocyte ratio (PLR) and modified Glasgow prognostic score (mGPS) were studied ^15^. In patients with locally advanced and metastatic disease, the CRP/alb ratio was an independent factor of poor survival ^15^. Another retrospective study evaluating CA19-9, CEA, CRP, LDH and bilirubin levels in locally advanced and metastatic pancreatic cancer patients treated with chemotherapy showed an independent prognostic significance for overall survival only for CA 19-9 decline during treatment ^16^. Other studies have evaluated risk factors for thromboembolic events in pancreatic cancer patients and more generally in patients with cancer ^17^ (see also Box 2). The Khorana score, developed more than ten years ago, is widely used to estimate venous thromboembolic risk in the population of cancer patients ^18^; it integrates standard laboratory parameters (platelet count, hemoglobin, leukocyte count), body mass index (BMI) and the cancer site (with pancreatic cancer and gastric cancer classified as very high risk). Still, its performance was questioned in a retrospective cohort of pancreatic cancer patients ^19^ and in a prospective cohort study of patients with different cancer types, among them 109 with pancreatic cancer ^17^. The clinical association of PDAC, sarcopenia/cachexia and thromboembolism is well-described ^11^, but still not understood in its pathophysiology ^20^. Within the SASKit study we aim to identify biomarkers and molecular mechanisms contributing to this clinical association, by investigating their relation to clinically relevant outcomes.

### Ischemic stroke, prevalence and outcomes

Ischemic stroke (IS) occurs in the German population with an incidence of 236 per 100,000 per year ^21^. The mean age of acute stroke patients is 73-74 years, with more than 80% of patients being over 60 years old. After a first stroke, nearly 5% of patients suffer a second stroke within a year. Mortality after IS is about 12% within one year and about 30% within five years ^21^. Mild to moderately disabled stroke survivors showed an elevated prevalence of sarcopenia >6 months after onset of stroke compared with non-stroke individuals (13.2% vs 5.3%) ^22^. The mechanisms underlying sarcopenia include loss of muscle mass, reduction of fibre cross-sectional area and increased intramuscular fat deposition occurring between 3 weeks and 6 months after stroke in both paretic and non-paretic limbs ^23^. Comorbid, or subsequent cancer may facilitate sarcopenia after IS. A US nationwide inpatient sample study reported that 10% of hospitalized IS patients have comorbid cancer, 16% of them with gastrointestinal cancer and 1% with PDAC, and that this association may be on the rise ^24^. Additionally, within two years after IS, another 2% to 4% of patients receive a new cancer diagnosis ^25-27^. Within the SASKit study we aim to identify biomarkers to predict outcome after IS in terms of general health state (i.e. sarcopenia, deterioration of clinical performance, cognitive functioning, frailty) and quality of life, as well as (co-)morbidity, as we do for the PDAC cohort.

### Ischemic stroke, known biomarkers and clinical scores

In an early study of 956 patients with acute IS, determinants of long-term mortality were age, obesity, cardiac arrhythmias, diabetes mellitus, coronary heart disease and organic brain syndrome at discharge from hospital; interestingly, hypercholesterolaemia and smoking did not affect long-term outcome ^28^. More recent studies uniformly identified age and stroke severity, usually assessed on the NIHSS or similar scales, as biomarkers of long-term functional outcome and mortality after stroke ^29 30^. Fibrinogen has been related to long-term outcome after stroke ^31 32^. There have been conflicting data on the predictive value of serum bilirubin levels on the long term risk of cardiovascular disease. While some studies are in favor of a predictive value (e.g.: ^33-35^), others are not (e.g.: ^36^). Also, CRP levels have been reported to impact the functional long-term outcome after IS ^37^, and early neurological deterioration after IS has been related to decreasing albumin levels, elevated CRP and fibrinogen levels ^38^. Potential biomarkers for occult cancer in IS patients include elevated D-dimers, fibrinogen, and CRP; infarction in multiple vascular territories; and poor nutritional status ^39^. Interestingly, IS patients with elevation of at least two of the following coagulation-related serum markers, that is, D-dimer, prothrombin fragment 1.2, thrombin-antithrombin complex and fibrin monomer, in the post-acute phase of stroke, were more likely to have occult cancer or recurrent stroke during follow-up for 1.4±0.8 years ^40^. In another study of acute IS patients, high D-dimer levels at admission were independently associated with recurrent stroke and all-cause mortality during follow-up for up to 3 years ^41^. These findings underpin the idea of shared risk factors for unfavorable outcomes in IS as well as cancer and they suggest that there may be coagulation-related biomarkers indicating an early stage of carcinogenesis or stroke (see also Box 2). Nevertheless, the clinical biomarkers that currently exist for predicting outcome are limited in their performance and clinical utility, and there is a need to overcome the limitations of current predictive models ^42^.

## Methods

The presentation is based on the reporting recommendations for tumor marker prognostic studies (REMARK), that is, items (1) – (11) of the REMARK checklist ^74^.

### Study design

The SASKit (“Senescence-Associated Systems diagnostics Kit for cancer and stroke”) study is designed as a prospective, observational, cohort study to identify biomarkers for disease deterioration in patients with PDAC or with IS and, specifically, for the (co-)morbidities of these diseases including vascular events and sarcopenia following the diagnosis of PDAC as well as cancer and cognitive decline following IS. All patients will be treated for their diseases in accordance with current guidelines or therapy standards and at the physician’s discretion. Due to the observational study design, regular treatment of the patient is not affected apart from sampling blood (20 to 80 ml at up to 7 time-points over the next years). Assessment of disease deterioration will be based on standardized clinical performance measurements, and patient reported outcomes based on questionnaires (see below for details). Additionally, data from clinical charts and information from the general practitioner will be collected. The SASKit study is divided into two subtrials with a common control group, both featuring essentially the same outcomes, predictor measurements and data analysis approaches.

### Characteristics of participants (patients and controls)

In the first subtrial (PDAC-subtrial), patients with an initial diagnosis of PDAC in locally advanced or metastatic stage without previous systemic therapy will be considered for enrollment, whereas patients with a (thromboembolic) IS of the supratentorial brain region within the past 5 to 10 days, with a definitive brain infarction volume >10 ml in an assessment by magnetic resonance imaging (MRI) will be considered for the second subtrial (IS-subtrial). Except for some explorative analyses, the subtrials will be analyzed separately.

Within both subtrials, eligible as controls are those without PDAC or IS and with no other malignant disease or other (hemorrhagic) stroke during the past two years. Potential controls will be recruited from persons who have lived in the same household as the patient within the last 2 years, have a maximum age difference of 12 years and are neither brothers nor sisters (i.e. spouses, second-degree relatives or friends). The controls are selected so that the age and gender structure approximately reflects the age and gender distribution of the patients. Therefore, the age and gender of the patients will be continuously recorded, and the controls selected in such a way that their frequency distribution of gender at any time corresponds approximately to that of the currently recruited patients.

The following criteria lead to exclusion from participation in the study for both patients and controls, *at time of recruitment*:

- previous or current medical tumor therapy
- other cancer within the past 2 years
- previous stroke with persistent deficit
- myocardial infarction within the past 2 years
- therapeutic anticoagulation within the past 2 years for longer than 1 month
- pre-existing dementia
- chronic heart failure stage NYHA IV
- terminal renal insufficiency with hemodialysis
- known HIV infection
- known active hepatitis C
- pregnancy
- age < 18 years.

Both subtrials will be implemented according to the same standardized protocol. After written informed consent of each participant, patients and controls will be followed up at 3, 12, 24, 36 and 48 months after their inclusion in the trial, whenever possible. The PDAC-subtrial includes an additional time-point for examinations at 6 months after inclusion, given that mortality due to PDAC is expected to be accelerated as compared to IS.

The study is expected to start in the second quarter of 2020 and will finish with the last participant’s follow up at 48 months. Until that time, we expect that 50 PDAC patients, 50 IS patients, and 50 controls participated in the trial. The study will be conducted at the Rostock University Medical Center (UMR), Germany at Clinic III - Hematology, Oncology, Palliative Medicine and at the Department of Neurology; the institutions of the other co-authors are supporting the study in a variety of ways. The study protocol has been approved by the ethics committee of the UMR. The study is registered at German Clinical Trials Register (DRKS00021184) and will be conducted following ICH-GCP.

### General health- and disease-related and demographic data

General data of the study participants will be recorded at the beginning of the study (“month 0”) and consist of the following: age, sex, BMI, temperature, blood pressure, heart rate (ECG). Furthermore, through interviews the following additional data will be recorded: vascular risk factors (arterial hypertension, diabetes, hyperlipidaemia, smoking habits), history of vascular events (stroke, myocardial infarction, venous or arterial thromboembolism), atrial fibrillation, history of cancer, current medication, surgery or blood transfusions in the past three months and vascular or cancerous events affecting any first degree relatives. These data may provide influential factors for explorative analyses, or be employed to interpret and discuss the results of the study.

### Blood sampling

Blood sampling will be done in a standardized fashion, that is, fasting and between 8 and 10 am, for all assays. Routine blood parameters will be recorded at the time-points described above (months 0 to 48). These consist of differential blood count, INR (International normalized ratio of prothrombin time), partial thromboplastin time, D-dimers, fibrinogen, factor XII, albumin, bilirubin, high-sensitive CRP, CA19-9, cholesterol, and HbA1c.

Experimental blood analysis (PAI-1 and omics) will be done for patients at month 0 in case of PDAC, at month 0 or at month 3 in case of stroke (where the 3-month time point is taken if it reflects a better state of the patient as described by the NIHSS), and furthermore at month 3 in case of PDAC, and at month 12 in case of stroke. For controls, the experimental blood analysis will be carried out at month 0 and at month 12, assuming that for these, data do not change much in the 3 months after baseline. The justification for taking the better state in case of stroke is the maximization of differences with the 12 months follow-up data. In terms of practicality (being able to calculate a biomarker signature sooner), however, the state at month 0 should be selected for all stroke patients. Since the blood sample will be taken pre-processed and frozen at month 0 in all cases, we are in principle able to perform the experimental blood analysis for all stroke patients at month 0, and we can do this analysis in retrospect if deemed necessary. We also take blood of PDAC patients at month 12, to have the option to do an experimental blood analysis if deemed useful. In the following we will refer to the *baseline* time-point (month 0, or month 3 in cases of stroke patients that improved) and the *landmark* time-point (month 3 for PDAC patients and month 12 for stroke patients and controls). The experimental blood analysis is done earlier for PDAC because of high expected mortality within the first year.

The experimental blood analysis includes PAI-1 (see Box 2) as well as high-throughput (omics) analyses, that is, transcriptomics and proteomics analysis in T-cells and proteomics of serum. T cells are of interest because these were reported to carry the strongest signal with respect to cellular senescence, based on the marker p16 ^75^. We intend to measure gelsolin and osteopontin as well, provided that sufficiently standardized assays become available in due time; the blood collected for this measurement shall otherwise be used to measure cytokines/chemokines such as IL6, IL8 and TNFC, which are part of the SASP, by ELISA assays. At time of writing, we do not yet have reliable estimates on the amount of blood cells still available for measuring protein expression, so an antibody-based protein array (in case of low amounts), or mass spectrometry (in case of sufficiently high amounts) will be used alternatively. For the blood serum, we intend to use the same protein measurement method. In the default case of a protein array, we plan to use the novel but dedicated “Senescence Associated Secretory Phenotype (SASP) Antibody Sampler Kit” (consisting of approx. 10 SASP-related proteins being measured; Cell Signaling Technology) for both cellular and serum proteomics. Further exploratory molecular analyses not (yet) funded but permitted based on the ethics approval include the following: single-cell analyses of blood, methylation assays for calculating epigenetic clocks ^76^, genetics by SNP array or whole-genome sequencing, and telomere length. A separate ethics approval was granted for an optional skin biopsy; skin microbiome analyses are planned as well.

Blood sample processing for the experimental analysis will be performed according to standard operating procedures (SOP) at the research laboratory of Clinic III - Hematology, Oncology, Palliative Medicine. The procedures include flow cytometric control of the sampling quality including distribution of cell types and vitality as performed in routine diagnostics. Isolation of peripheral blood mononuclear cells (PBMCs) will also be performed following the SOP used by the laboratory in routine diagnostics. T-Cell separation will be performed according to an established work flow based on magnetic bead purification via Miltenyi MACS following manufacturer’s instructions. T cell fraction purity as well as vitality will then be verified by flow cytometric analyses as described above. Nucleic acid isolation as well as protein isolation will be further performed according to the SOP of the research laboratory performed using column separation (Qiagen, Hilden Germany). RNA integrity values (RIN) will be analysed using an Agilent Scientific Instruments Bioanalyzer as instructed by the manufacturer. RIN values above 6 will qualify for RNAseq or Clariom D Array analyses; for RNAseq average reads per sample will be set at approx. 40 × 10e6.

### Clinical performance measurements and patient-reported outcomes

At baseline and at each follow-up, handgrip strength (“grip strength” for short) is measured using a digital hand dynamometer (Jamar Plus). The test is performed while sitting comfortably, shoulder adducted, elbow placed on the tabletop and flexed to 90 degrees, with the forearm and wrist in a neutral position^77^. The highest value of three measurements of maximal isometric contraction of the dominant hand, or if paralyzed due to IS, contraction of the unaffected hand, is documented in kg. Further, the following clinical performance measurements are evaluated by the study physician or study nurse according to standard protocols: ECOG Performance Status (ECOG PS)^78^, modified Rankin Scale (mRS)^79^, Canadian Study on Health & Aging Clinical Frailty Scale (CSHA-CFS)^80^, NIH-Stroke Scale (NIHSS)^81^, Montreal Cognitive Assessment (MOCA)^82^. All raters are certified for the applicable scores (mRS, NIHSS, MOCA). Patient-reported outcomes (measured by questionnaires) are the following: EQ-5D-5L and EQ-VAS (generic evaluation of QoL in 5 domains and overall on a visual analog scale)^83^, HADS-D (evaluation of anxiety and depression)^84^, WHODAS 2.0 (WHO Disability Assessment Schedule)^85^, and, for patients with PDAC, FACIT-Pal (evaluating QoL with focus on palliative symptoms and needs)^86^,^87^. All questionnaires are administered following the suppliers’ instructions.

### Follow up data

Apart from the clinical and patient-reported outcomes, further follow-up data are BMI, temperature, blood pressure, heart rate (ECG), atrial fibrillation, current medication, tumor treatment, comorbidity (any vascular or cancer event), hospital admissions or palliative care. Additionally, based on clinical charts and information from the general practitioner, we will record medication, (co-)morbidity and mortality. Just like the general health- and disease-related and demographic data recorded at time of recruitment, these data may provide influential factors for explorative analyses, or be employed to interpret and discuss the results of the study.

### Endpoints

In both subtrials, the primary endpoint is a composite measure of “disease deterioration” defined as the first occurrence within a follow-up interval of at least one of the following.

a. Sarcopenia, measured by grip strength less than 27 kg for males and less than 16 kg for females (according to the revised European consensus, EWGSOP2, ^1^).
b. Deterioration of clinical performance, that is, of the ECOG PS by at least two points (PDAC-subtrial), or of the mRS by at least one point (IS-subtrial).
c. Deterioration of QoL, described as a reduction of the EQ-5D-5L by at least 0.07 in the index score, ***and*** deterioration of at least 7 points in the EQ-VAS (ranging from 0-100).

Deterioration will be considered between baseline (month 0) and the respective follow-up investigation. As described above, for patients with IS who have improved their condition (measured by NIHSS) within the first 3 months, this time point (month 3) will be used as a baseline instead. Item (a) is the deterioration from “no sarcopenia” to “probable sarcopenia” as defined by current consensus^1^. Grip strength has been widely used for assessing muscle strength, which is currently used as the most reliable measure of muscle function, loss of which indicating sarcopenia^1^. ECOG PS is established in describing the general condition of patients with cancer, whereas mRS is established in patients with stroke. Death is reflected by both scores as ECOG PS of 5 or mRS of 6, and it will always consider death from any cause. The EQ-5D-5L evaluates QoL in five dimensions (mobility, self-care, usual activity, pain/discomfort, and anxiety/depression), all relevant for patients with PDAC and IS. Furthermore, it is a generic score so that results will be comparable for different diseases (as recently described in patients with stroke^88^) and for the general population^89^). Even though disease-specific scores might evaluate symptom burden in even more detail, the EQ-5D-5L was recently shown to be comparable to QoL scores developed specifically for pulmonary embolism and deep vein thrombosis (that is, PEmb-QoL, VEINES-QOL/Sym and PACT-Q2) in terms of acceptability, validity and responsiveness^90^. A clinical deterioration in EQ-5D-5L is described as a minimal important difference in the range from 0.07 to 0.09 index points and in VAS from 7 to 10^91^which is the basis for the definition of item (c). Controls reach their endpoint by the same definition as the subcohort for which they serve as control; in any integrative analysis of both subtrials, a deterioration of the mRS by at least one point will be used as the criterion (instead of ECOG PS), because stroke patients in general have a slower deterioration than PDAC patients, and controls naturally have the slowest expected deterioration.

The primary composite endpoint and all secondary endpoints will be evaluated in a first analysis, based on data obtained until summer 2021, and in a second analysis, based on data obtained until summer 2023, and in a third analysis at the end of the study. The second analysis may be delayed until data of 90% of the study participants are available (at least including the month 12 follow up) and it may then constitute the “main” analysis of the study.

The following secondary endpoints are evaluated:

- each component of the primary endpoint (separately);
- occurrence of disease-specific (co-)morbidities, as follows

- new vascular events (stroke, myocardial infarction, venous or arterial thromboembolism), specifically in patients with PDAC;
- new cancer, specifically in patients with IS;
- probable sarcopenia (based on grip strength);
- cognitive decline (deterioration of MOCA by 3 points from best value at baseline);
- frailty, defined as a CSHA-CFS level of 6, 7, or 8;
- all-cause mortality.

Further, a sum-score summarizing all measurements of phenotypic variables (grip strength, clinical performance measurements, comorbid events, mortality) will be considered as a surrogate for “aging”, normalizing all continuous-scaled components in order to obtain a common scale with an average of zero and standard deviation of one. The components of the sum-score will all be given equal weight.

### Predictors

While all phenotypic features (grip strength, clinical performance, patient reported outcomes, comorbid events, mortality) are contributing to the definition of endpoints (as dependent variables/parameters), all routine and experimental blood features (PAI-1, omics) are considered to be potential predictors; these are also called the independent variables/parameters. This delineation is justified by (a) the paradigm that (clinical) relevance is tied to high-level phenotypes describing health and survival, specifically including QoL^2^, and (b) the goal of developing a “senescence-associated systems diagnostics kit” that includes a careful selection of biomarkers contributing, as much as possible, also to molecular-mechanistic insights into PDAC, IS and their (co-)morbidity, which we hypothesize to be related to cellular senescence and aging. Age and gender will be included as mandatory covariates (also termed confounders, that is, predictors which we do not aim to explore, or which we wish to improve upon) in all statistical models. Further covariates are smoking, the baseline NIHSS score in case of IS, as well as locally-advanced vs metastatic PDAC and modality of treatment in case of PDAC. As described, the successful predictors identified by our study, following the statistical analyses outlined below, are called biomarkers; we wish to stress that these are only *candidates* for the ultimate goal of *clinically validated biomarkers*; in particular, they still need to be validated in further studies (based, e.g., on other cohorts). A set of biomarkers is also called a biomarker signature.

### Blinding and pseudonymization

No blinding will be done during the study. However, the primary composite endpoint will be documented without subjective influence due to standardized definitions. Thus, detection bias will be kept at a minimal extent. Furthermore, information bias will be minimized as we will use simple measurements, which are applied in daily practice or are self-reported and easy to perform (e.g. EQ-5D-5L). The rigorous inclusion of all eligible patients within the recruitment period will help to minimize selection bias. All patient data are pseudonymized to all investigators except for the attending physician and study nurse. Since all major data analyses are based on known information about the outcomes (e.g., supervised machine learning with cross-validation), the data analysis will also be performed based on the pseudonymized data. Protection of personal and clinical data of all patients and controls will follow all relevant legal regulations.

### Sample size

No formal sample size calculation was performed a-priori for this observational study. The prevalence of PDAC combined with the requirement to complete the study within a reasonable timeframe implied a target of 50 patients per group (PDAC, IS and control group). Nevertheless, a power analysis revealed that a sample size of 50 patients will have 80% power to detect a significant difference by a non-parametric Wilcoxon statistic between an AUC of 0.75 for a particular biomarker signature compared to the null hypothesis value of 0.5 at a significance level of 5% under the assumption that about three times as many patients will reach the primary endpoint, compared to patients who will not reach the primary endpoint^92^.

### Data Analysis Plan

#### General considerations

The guiding criteria for biomarker identification in the SASKit study are the maximization of the predictive signal, clinical relevance/utility, biomedical/molecular/clinical interpretability, and practicality/cost. Given the relatively low number of participants in this in-depth study, to maximize the signal for the endpoints and predictors given as outlined above, we must aim to use all available information. Regarding endpoints, whenever possible, we thus wish to consider the (censored) time-to-event information inherent in the baseline and follow-up examinations, and in the mortality data. The primary endpoint was defined to integrate expected clinical utility and maximum signal. In defining the (secondary) endpoints, we considered an array of clinically relevant single endpoints as well as a sum-score of all phenotypic measurements; we hypothesize that the latter carries the largest amount of signal. Given the small sample, we cannot set aside an extra validation dataset. (For the predictors considered to be covariates/confounders, please see the section on “Predictors”, above.)

### Data quality assessment and cleaning

The need for (and the amount of) data cleaning cannot easily be estimated beforehand; we plan to follow the MarkAGE guidelines ^93^ to deal with missing values, and to detect and rectify outliers and batch artefacts.

### Predictor/Feature integration

Regarding predictors (features), we first need to remember that we measure at baseline (at months 0 or 3) and at one landmark (main followup, that is, at months 3 or 12). While use of baseline features is unrestricted, use of landmark features is, of course, restricted to predict outcomes after the landmark. Further, we need to handle the high dimensionality of the omics features. Here, upfront feature integration, e.g., by averaging measurements as described below, is considered preferable specifically for the high-dimensional omics data, for the following reasons.

1. A small feature space allows for an easier understanding and interpretation, see, e.g., ^94^.
2. Integrated features can be used as input for both the standard biostatistics and the standard machine learning parts of the analysis.
3. Use of few features is more time-tested than newer methods featuring the joint calculation of the prediction model and the selection of the features, albeit the latter are quite often claimed to be superior by their developers.
4. Naturally, feature integration avoids multicollinearity and overfitting, and multiple testing is less of an issue. This counters the “curse of dimensionality” and “de-noises” the data towards better prediction performance ^94 95^.
5. Feature integration allows the handling of feature heterogeneity, which in our case refers to routine blood measurements as well as various omics data types.
6. In the *explorative* analyses, systems biology modelling and the parallelogram approach are both supposed to deliver further small sets of integrated, highly informative features, which may, e.g., dominate systems behaviour, or which are believed to translate well from animal models to humans (see below).

While most features will be available for the baseline and the landmark time-point, utilizing baseline data is clinically more useful, simply because the prediction for the endpoint is available much earlier. Nevertheless, in the explorative analyses, we will investigate the predictive power of *changes* in feature measurements from baseline to landmark, given that such changes may be more informative about future disease deterioration (and other endpoints) than just baseline values.

#### Specific omics data feature integration

Notably, we face a heterogeneous “multi-view” dataset, usually referred to as “multi-omics”. Our feature integration approach (see above) is also known as a “late integration” type of analysis, implying that measurements for different omics data types are reduced early on to activation scores for pathways or subnetworks that are then integrated at a “late” level. To calculate the activation scores for subnetworks, we use, by default, the ExprEssence/FocusHeuristics linkscore ^96 97^, taking the links (gene/protein interactions) from a functional interaction network defaulting to STRING. Our experience with the *linkscore* motivates us to include this method as one of the approaches proposed for feature integration in the following, influencing the calculation of up to 10 features on which the standard biostatistics and machine learning shall be based. Specifically, we take the average expression measurement for all patients (as a list of expression values, one per gene) and the average for all controls (as a list of expression values, one per gene) to calculate a *linkscore* for each STRING interaction, and assemble a “condensed” network including all interactions with a *linkscore* in that percentile for which the 50 highest-scoring interactions are shown. These interactions form subnetworks. We then take the average *linkscore* for each subnetwork as the subnetwork activation score. Alternative methods such as *keypathwayminer* will be used in the exploratory analyses, see below. For the pathways (such as KEGG), we will calculate pathway activation scores using Gene Set Variation Analysis (GSVA) ^98^. This method calculates pathway activation scores from expression data, is suited for use with microarray as well as RNAseq data and performed strongly in a recent benchmarking analysis ^99^. The GSVA-based pathway activation scores can subsequently be compared between patients and controls in the same way as normal gene expression data, calculating, for each pathway, a fold-change of the pathway activation scores between patients and controls. Here, we average over all patients and over all controls, respectively, using the *limma* R package and adjusting for age and gender of the individual patient/control pathway activation. An example of this approach is given in the GSVA publication, where differential pathway activation was identified between acute lymphoblastic lymphoma and mixed-lineage lymphoma ^98^. The major downside of feature integration may be information loss; subsequent statistical and machine-learning-based analyses receive only a tiny fraction of the amount of information that is available in total.

Gene expression data (transcriptomics) will be our preferred omics data type. Nevertheless, proteins are closer to the phenotype than transcripts, so we wish to not ignore these. Therefore, we prepare to deal with both kinds of proteome data that we may expect (see “Experimental blood analyses”, above), as follows.

1. Large-scale data, likely based on mass spectrometry, in the order of hundreds or more proteins that can be identified and measured in all the conditions investigated differentially.
2. Small-scale data, likely based on antibody arrays, in the order of tens or less.

Except for the raw data preprocessing depending on the platform, once log-fold changes describing differential expression are established, we thus expect to handle the large-scale proteome data essentially the same as the transcriptomics data, and the small-scale proteome data similarly to the blood routine data, for cells and serum alike. Overall, the omics data are expected to come along three main coordinates, that is,

1. as blood cell transcriptomics and proteomics as well as serum proteomics;
2. longitudinal in time (for baseline and landmark); and
3. for PDAC, IS and control.

All coordinates can be exploited for differential analyses, even though the PDAC and IS data will be analyzed separately except for some integrative *explorative* analyses (see below). In the *explorative* analyses, the *longitudinal* transcriptomics of the patients and controls will also be analyzed together, see below. For the standard biostatistics and machine learning analyses, we plan to employ 5 approaches to feature integration, each yielding a shortlist of 5 integrated features, as follows.

1. **(*5 features*)** A first shortlist of features will consist of the following expert selection from the routine blood measurements (incl. PAI-1): *neutrophil-lymphocyte-ratio, fibrinogen, high-sensitive C-reactive protein, albumin and PAI-1*.
2. **(*5 features*)** For the cellular gene expression measurements, we use ExprEssence/FocusHeuristics (see above) to calculate the top-5 subnetworks scoring highest.
3. **(*5 features*)** Again for the cellular gene expression measurements, we use GSVA (see above) to calculate the top-5 most strongly changing pathways as features.
4. + 5) **(*10 features*)**
  a. In case of dealing with large-scale serum proteomics data, we proceed as in (2) + (3);
  b. In case of dealing with small-scale serum proteomics data, we proceed as follows:
    i. if the number of features measured successfully is in the order of 10, we refrain from any processing;
    ii. if the number of features is in the order of around 10-100, we select the 10 features with the smallest p-values indicating differences between the mean values of patient and control, based on a t-test.

For genomic features as per (2), the feature measurements for an individual patient or control will then be the average linkscores of the 5 selected subnetworks. For genomic features as per (3), the feature measurements for each patient/control will be the GSVA scores of the 5 selected pathways. By construction, we expect the resulting features to reflect the up/downregulation of disease-related transcripts/proteins or pathways/subnetworks. Using the GSVA-based integrated features as input to the biostatistical analyses employing Cox proportional hazard models, we are in fact closely following the “Survival analysis in ovarian carcinoma” example as described in the GSVA publication ^98^. Regarding the expert selection from the routine blood measurements, we are aware that some of these features may be considered to have an almost trivial relationship to outcome prediction for the diseases we study; e.g. fibrinogen may correlate strongly with the size of the stroke-damaged brain area and may thus be considered a covariate. However, to our knowledge, none of these features are validated clinical biomarkers, and it is quite possible that a combination of simple biomarkers is key to the best possible prediction. We selected the *neutrophil-lymphocyte*-ratio specifically because it is cheap to measure; it is, however, like many other blood-based features, easily influenced by acute infection.

#### Exploratory feature integration

Apart from the FocusHeuristics/ExprEssence *linkscore*, we employ alternatives such as *keypathwayminer* ^100^. Further, we calculate pathway activation scores for the following senescence-related KEGG pathways, which include PAI-1 (see the Introduction) but do not refer to a specific disease, as of February 2020: *Cellular senescence, HIF-1 signaling pathway, p53 signaling pathway, Apelin signaling pathway, Hippo signaling pathway, Complement and coagulation cascades*. “Early integration” by, e.g., first averaging transcript and protein expression on a single-gene basis, is also planned.

#### Choice of data analysis methods for biomarker discovery

We will consider two main approaches of data analysis, one motivated by statistical methods, the other by machine learning approaches. While this delineation may ultimately be meaningless, we consider that regression is the core ingredient of the former, while supervised learning characterizes the latter. We will apply “standard” methods (mostly in biostatistics) and explore novel approaches (mostly in machine learning; preserving signal implies a focus on supervised approaches in this case). Data analysis for biomarker discovery trials in a *clinical* setting is usually described with a biostatisticians’ mindset, who also developed methods to cope with the high dimensionality of omics data (see below). On the other hand, the challenges of omics data also spurred the recent publication of many methods adopting machine learning, which however did not yet make it into clinical trial analysis routine, but which we wish to test (see below). We will focus on methods readily available in SAS or as R packages. Notably, the correct choice of method depends in part on known unknowns such as the strength of the signal (incl. the amount of missing data) in the routine blood measurements and the omics.

#### Prediction model quality measures

Unlike intervention trials with their highly standardized aim of establishing a statistically significant superiority (or non-inferiority) of one intervention compared to another (or to standard of care), observational biomarker trials are a more recent development with fewer precisely quantified criteria of success, and a stronger need to consider the effect size: even if a biomarker signature enables a significant improvement in predicting an outcome, raising the accuracy of the prediction, say, from 70% to 75% may not be clinically meaningful, depending on prevalence of the condition to be predicted, the cost of the biomarker measurement, etc. We thus aim to identify biomarkers making a maximum of *difference* in prediction accuracy, if we are able to compare to established scores (see also below). For the biostatistics part, the concordance statistics (c-index) will be used as an overall measure of predictive accuracy, and time-dependent ROC curves and AUC will be used to summarize the predictive accuracy at different cut-off points in time. For the machine learning part, the cross-validated accuracy and AUC/c-index, following ^94^, are used, and to take care of a potential Simpson’s paradox we will either analyse the data stratified by gender, or we will add such an analysis and check for consistency. More generally, to investigate the role of confounders (and, if necessary, to correct for these) in the machine learning part, we wish to use the permutation technique described in ^101^. We expect that we can identify a set of biomarkers that affords an accuracy of 75% or more or an AUC of 0.75 or more in correctly predicting the primary endpoint with a precision of +/- 12% ^102^. This estimate of precision is based on half the width of a 95% confidence interval (CI) for a probability of 75%, by extension of Table 6 of ^102^, which shows precision up to a sample size of N=30.

#### Standard biostatistical analyses

A Cox proportional hazards regression model adjusted for age and gender will be used to estimate the hazard ratio (HR) and corresponding 95% CI to predict the primary composite endpoint separately within the PDAC cohort and IS cohort. The 5 shortlists of 5 features (see above) will be providing the canonical predictors, analyzed together. For selection of the most important features that might be related to the primary endpoint we will use a procedure proposed by Sauerbrei et al. ^103^, as follows. First, 100 bootstrap samples will be generated. Then, a multivariate Cox proportional hazards regression model with backward elimination with selection level of 0.05 will be fitted to each replication of the original data set. In a second step features with a relative selection frequency of 30% or less over all bootstrap samples will be eliminated. In a third step each feature Xi for which the hypothesis of independence in combination with a feature Xj can be rejected will be eliminated if Xi is less important when Xj is included in the model, or if it does not gain importance when Xj is excluded from the model. All remaining features will be included in the final model. Graphical and numerical methods will be performed to establish the validity of the proportionality assumption ^104^ in the final model. Results will be reported as p-values, HRs and corresponding 95%-CIs. A p-value of p ≤0.05 will be interpreted as indicating statistical significance. From the final model a risk score will be calculated by multiplying the individual feature measurement of a patient with the estimated regression coefficient of each feature. The c-index will be used as an overall measure of predictive accuracy of the resulting score, a time-dependent ROC curve and AUC will be used to summarize the predictive accuracy of the score at specific times. All secondary endpoints will be evaluated using the same approach as for the primary endpoint except for the sum-score used as a surrogate for “aging”. For this endpoint, a linear mixed effects model with random intercept and spatial power covariance structure will be fitted to the data to estimate the progression of “aging”. The covariance structure is chosen to reflect the unequal intervals of follow up investigations. Model assumptions and model fit will be checked by visual inspection of residuals, and influence diagnostics. Missing values will be taken into account by a likelihood-based approach within the framework of mixed linear models with the assumption that missing values occur at random. Results will be reported as p-value assessed at a level of significance of 5% accompanied by the value of the test statistic and degrees of freedom. In addition, 95% CIs for the progression (slope) will be provided.

#### Additional exploratory biostatistical analyses

Again, the primary composite endpoint as well as all secondary endpoints will be evaluated separately within the PDAC cohort and IS cohort of the respective sub-trials. In a first approach, univariate Cox proportional hazard models adjusted for age and gender will be calculated for each omics feature (R package *survival*) using a cut-off of 0.05 on the false discovery rate. In a second approach, all omics features will be simultaneously considered in a multivariate Cox model, adjusted for age and gender. Towards this aim, a component-wise likelihood-based boosting algorithm proposed by Binder and Schumacher 2008^105^ (R package *CoxBoost*) will be used to develop a biomarker signature.

#### Standard machine learning

For the machine learning part, the primary outcome and all secondary outcomes give rise to an assignment of predictor/feature lists to survival times, one such list per study participant, for which biomarkers are then learned in a supervised fashion. As described, in the standard analyses, feature integration (see above) will precede the actual calculation of the model (“deep” learning approaches that take in “all” features are part of the *exploratory* analyses, see below). In the same way as the standard biostatistics analyses, the same 5 shortlists of 5 features each (see above) will be providing the canonical predictors, analyzed together. Exploiting time-to-event information, we will employ random survival forests (RSF) as described by ^106^with the following advantages.

1. RSF can now be considered a time-tested approach, and it was the subject of a recent extensive review ^65^ and of a systematic comparison with LASSO approaches in the case without feature selection (^107^, see their Table 7 for its competitive performance which is not reflected in their abstract).
2. RSF can also work on essentially all features, without a preceding feature integration/selection step, and then be compared, in the explorative machine learning analyses described below, to survival support vector machines (SSVM) and to a novel method Path2Surv that “conjointly” performs feature selection and model training, see ^94^.
3. RSF was recently compared to Cox-nnet ^108^, a neural network approach which we consider as very promising for the *exploratory* part, see also below.
4. RSF offers a considerable degree of interpretability, given that RSFs are derived from decision trees.
5. RSF is considered “completely data driven and thus independent of model assumptions” and “in case of high dimensional data, limitations of univariate regression approaches such as overfitting, unreliable estimation of regression coefficients, inflated standard errors or convergence problems do not apply”^65^.

In the machine learning part, we calculate accuracy and AUC/c-index using cross-validation to make the best use of our limited sample size, following the setup of ^94^ and ^107^ (who, however, set aside separate validation datasets).

#### Additional exploratory machine learning

Apart from the more time-tested standard machine learning described above, we will also explore methods that were proposed recently, for which it is less straightforward to tell whether these methods are fit-for-purpose in our case, even though they are usually claimed to be superior by their developers based on some test/validation data sets. Specifically, as mentioned above, we expect to test Path2Surv and SSVM ^94^ as well as Cox-nnet ^108^ (without prior feature integration); the latter in particular promises a high degree of interpretability. We further explore CNet (employing the censored-data variant), for interpretable biomarkers. We also plan to employ the PASNet ^109^, SurvivalNet ^110^ and SVRc ^70^ packages. The longitudinal transcriptomics of the patients and the controls may also be analyzed integratively based on the “optimal discovery procedure” ^111^, considering, however, that landmark feature data can only be used to predict events after the landmark. Finally, we will map the differential omics data onto a human “healthspan pathway map” ^112^, that is, a set of clusters/pathways based on health-related genetic data that we assembled recently.

#### Explorative systems biology modelling, explorative parallelogram approach and transfer learning

As mentioned, systems biology modelling and parallelogram ^113 114^ extrapolation are supposed to deliver small sets of highly informative features, by contributing features that are dominating model behaviour or that are shown to translate from the SASKIt animal model data. Given the comparatively small number of study participants (but in-depth measurements), we also wish to explore “transfer learning”, which aims to utilize large amounts of public knowledge in the form of latent variables. Specifically, we plan to use, and wish to develop further, the Multiplier ^115^ approach motivated by the analysis of rare-disease data. Multiplier utilizes the RNASeq-based recount2 compendium, and apart from the functional network and pathway data that we use in the feature selection part, this compendium is expected to be our main source of biological knowledge that enters the calculations for biomarker discovery.

#### Miscellaneous exploratory approaches and discovery of diagnostic biomarkers

We will also use unsupervised machine learning to generate descriptive multi-omics correlation networks, as they were most recently employed by ^116^, there supplemented by linear mixed effects models using (un-)restricted maximum likelihood approaches; in this very recent biomarker discovery trial of similar design as ours, but with many more longitudinal omics measurement time-points than ours, we could not identify other biomarker discovery methods being used. If genetic data become available, we will include these in some analyses; specifically, we will investigate the added value of *expression quantitative trait loci* (eQTL) analyses. PDAC and IS data will be analyzed together in some integrative exploratory analyses. In that case, the occurence of specific endpoints will be evaluated according to the group membership (PDAC or IS). This means that in addition to the biomarker signature, a group variable, indicating PDAC or IS patients, will be included in the analysis, to assess the difference in the progression of the respective endpoints between PDAC and IS patients. We also wish to compare PDAC and IS patient data to data of healthy controls (adjusted for age and gender) by means of logistic regression models with the aim of identifying candidate biomarkers for the diagnosis of the respective disease; we then specifically investigate the association of these diagnostic biomarker candidates with cellular senescence and other aging-related processes (see also the next paragraph).

#### Further analyses, and comparison with existing biomarkers and biomarker signatures

Towards the end, we will investigate the overlap for the various biomarker identification approaches we employed, assuming that the most frequently found biomarkers may be the most robust and valid ones. Moreover, we will compare with existing biomarkers and signatures. Regarding the prediction of vascular events, we will specifically calculate the Khorana and related scores^17^ for comparison, and report the difference in performance. Further, for all biomarkers we find, we will check their association with cellular senescence, by manual inspection, literature investigation, comparison to CellAge^117^ and the SASP Atlas^50^ or by formal enrichment analyses if the number of biomarkers is sufficiently large to do this in a meaningful way. Also, in a final step, we plan to identify and filter out the biomarkers that are volatile in the controls. In addition, a comparison of the biomarker profiles before and after the co-morbid event is aimed for. Finally, for publicly available data of other trials with a sufficient overlap with our predictors, we will use these as validation datasets.

## Discussion

### Limitations

Arguably, the most serious limitation of the SASKit study is the low number of participants. We mentioned above that in the 4-year-time-frame of the entire study, at the Rostock University Medical Center we cannot expect to recruit many more than the 50 PDAC patients to be included in this study; we could recruit more stroke patients and more controls, but given the call for proposals that allowed this exploratory (not confirmatory) study to be applied for and funded, we considered that within a limited budget, in-depth omics characterization, animal models (to be detailed in a follow up publication) and a comprehensive data analysis plan including systems biology modelling were important aspects of our study that we did not want to exclude.

The two most obvious risks to the main goal of finding good biomarkers for the primary outcome based on the standard data analysis are the following. First, we found it hard to estimate the distribution of events as defined by the primary outcome; we cannot exclude that too many events take place already at the start of the study, or until the first follow-up, specifically in the PDAC subtrial, limiting the amount of information available to the subsequent time-to-event analyses. Then again, had we defined the primary outcome more conservatively, there would have been a chance that not enough events happen until the end of the study. Second, we could not identify role-model publications reporting results of biomarker explorations that made use of machine learning methods, except for, to some extent,^116^, so that we enter unknown territory to some degree. The two most obvious risks to our goal of investigating the role of cellular senescence in the (co-)morbidity of PDAC and IS could be an insufficient prevalence of co-morbid events, and the complex role of treatment in case of PDAC, where additional cellular senescence is most likely triggered by therapeutic intervention^118^. Then again, all molecular high-throughput analyses are essentially explorative and we are open to discovering biomarkers of disease that do *not relate* to any of our pre-specified hypotheses.

### Implications

We designed the SASKit study to synergistically deliver upon a couple of aims that we consider to be of relevance for specific disease prognosis and treatment as well as for primary, secondary and tertiary prevention. Employing clinical performance measurements and patient-reported outcomes, we aim for clinical relevance and we suggest that prognostic biomarker signatures for general health and QoL are perhaps more important than (progression-free) survival, although there is much more data about the latter than the former. Moreover, good disease treatment options are still lacking for PDAC as well as for stroke, and the more we find cellular senescence implicated in disease deterioration, at least in a subgroup of patients with a specific biomarker signature, the more confidently we can suggest, and further explore, seno-therapeutic interventions for these two diseases.

Notably, we are in the process of starting a parallel human study testing, in healthy elderly people, interventions into cellular senescence, based on *food* rich in seno-interventional compounds, and we expect that many aspects of the study design presented herein will be adopted in that parallel study. That study will also investigate aging- and senescence-related outcomes, and as such it can be seen as a test of a cautious yet potentially very effective approach to primary prevention; if the *diagnostic* biomarkers we find in the SASKit study relate to cellular senescence, this observation would constitute further evidence for (cautious) seno-interventions, moving towards a kind of universal approach of disease prevention by tackling fundamental aging-related processes (see Boxes 1 and 2).

Secondary prevention, aiming to reduce the impact of a disease that has already occurred, can ultimately be supported by the SASKit study, if we can demonstrate, and (in follow up studies) confirm, a distinctive role of cellular senescence (and/or other aging-related processes such as inflammation/inflammaging^119^) in disease deterioration as defined here. Finally, evidence for tertiary prevention by seno-therapeutic intervention, aiming to attenuate the impact of an ongoing disease, is also an option based on how accurate, relevant and specific our biomarkers will be.

Last but not least, we expect that the in-depth molecular analyses that we wish to conduct will provide mechanistic insights into the etiology of the diseases we study here, which we just see as models for the investigation of the fundamental role of aging in general and cellular senescence in particular in disease and dysfunction.

## Data Availability

N/A

## Abbreviations

AUC: Area Under the Curve
BMI: Body Mass Index
CA19-9: Carbohydrate Antigen
CEA: Carcinoembryonic antigen
CI: Confidence interval
CRP: C-reactive protein
ECOG: Eastern Cooperative Oncology Group
HR: Hazard ratio
INR: International normalized ratio
IS: Ischemic Stroke
LDH: Lactate dehydrogenase
NIHSS: NIH-Stroke Scale
NYHA: New York Heart Association
PDAC: Pancreatic Ductal Adenocarcinoma
PS: Performance status
QoL: Quality of Life
ROC: Receiver-Operator Characteristic
RSF: Random survival forests
SASKit: Senescence-Associated Systems diagnostics Kit for cancer and stroke
SASP: Senescence Associated Secretory Phenotype

## References

1. Cruz-Jentoft AJ, Bahat G, Bauer J, et al. Sarcopenia: revised European consensus on definition and diagnosis. Age Ageing 2019;48(1):16–31. doi: 10.1093/ageing/afy169 [published Online First: 2018/10/13]

2. Fuellen G, Jansen L, Cohen AA, et al. Health and Aging: Unifying Concepts, Scores, Biomarkers and Pathways. Aging and Disease 2019;10(4):883–900.

3. Collaborators GBDPC. The global, regional, and national burden of pancreatic cancer and its attributable risk factors in 195 countries and territories, 1990-2017: a systematic analysis for the Global Burden of Disease Study 2017. Lancet Gastroenterol Hepatol 2019;4(12):934–47. doi: 10.1016/S2468-1253(19)30347-4

4. Kleeff J, Korc M, Apte M, et al. Pancreatic cancer. Nat Rev Dis Primers 2016;2:16022. doi: 10.1038/nrdp.2016.22 [published Online First: 2016/05/10]

5. Llop E, p EG, Duran A, et al. Glycoprotein biomarkers for the detection of pancreatic ductal adenocarcinoma. World J Gastroenterol 2018;24(24):2537–54. doi: 10.3748/wjg.v24.i24.2537 [published Online First: 2018/07/03]

6. Carrato A, Falcone A, Ducreux M, et al. A Systematic Review of the Burden of Pancreatic Cancer in Europe: Real-World Impact on Survival, Quality of Life and Costs. J Gastrointest Cancer 2015;46(3):201–11. doi: 10.1007/s12029-015-9724-1 [published Online First: 2015/05/15]

7. Cruz-Jentoft AJ, Sayer AA. Sarcopenia. Lancet 2019;393(10191):2636–46. doi: 10.1016/S0140-6736(19)31138-9 [published Online First: 2019/06/07]

8. Taieb J, Pointet AL, Van Laethem JL, et al. What treatment in 2017 for inoperable pancreatic cancers? Ann Oncol 2017;28(7):1473–83. doi: 10.1093/annonc/mdx174 [published Online First: 2017/05/02]

9. Menapace LA, Peterson DR, Berry A, et al. Symptomatic and incidental thromboembolism are both associated with mortality in pancreatic cancer. Thromb Haemost 2011;106(2):371–8. doi: 10.1160/TH10-12-0789 [published Online First: 2011/06/30]

10. Grilz E, Posch F, Konigsbrugge O, et al. Association of Platelet-to-Lymphocyte Ratio and Neutrophil-to-Lymphocyte Ratio with the Risk of Thromboembolism and Mortality in Patients with Cancer. Thromb Haemost 2018;118(11):1875–84. doi: 10.1055/s-0038-1673401 [published Online First: 2018/10/09]

11. Bonnerot M, Humbertjean L, Mione G, et al. Cerebral ischemic events in patients with pancreatic cancer: A retrospective cohort study of 17 patients and a literature review. Medicine (Baltimore) 2016;95(26):e4009. doi: 10.1097/MD.0000000000004009 [published Online First: 2016/07/02]

12. Navi BB, Reiner AS, Kamel H, et al. Association between incident cancer and subsequent stroke. Ann Neurol 2015;77(2):291–300. doi: 10.1002/ana.24325 [published Online First: 2014/12/05]

13. Varki A. Trousseau’s syndrome: multiple definitions and multiple mechanisms. Blood 2007;110(6):1723–9. doi: 10.1182/blood-2006-10-053736 [published Online First: 2007/05/15]

14. Grilz E, Marosi C, Konigsbrugge O, et al. Association of complete blood count parameters, d-dimer, and soluble P-selectin with risk of arterial thromboembolism in patients with cancer. J Thromb Haemost 2019;17(8):1335–44. doi: 10.1111/jth.14484 [published Online First: 2019/05/18]

15. Liu Z, Jin K, Guo M, et al. Prognostic Value of the CRP/Alb Ratio, a Novel Inflammation-Based Score in Pancreatic Cancer. Ann Surg Oncol 2017;24(2):561–68. doi: 10.1245/s10434-016-5579-3 [published Online First: 2016/09/22]

16. Haas M, Heinemann V, Kullmann F, et al. Prognostic value of CA 19-9, CEA, CRP, LDH and bilirubin levels in locally advanced and metastatic pancreatic cancer: results from a multicenter, pooled analysis of patients receiving palliative chemotherapy. J Cancer Res Clin Oncol 2013;139(4):681–9. doi: 10.1007/s00432-012-1371-3

17. van Es N, Di Nisio M, Cesarman G, et al. Comparison of risk prediction scores for venous thromboembolism in cancer patients: a prospective cohort study. Haematologica 2017;102(9):1494–501. doi: 10.3324/haematol.2017.169060 [published Online First: 2017/05/28]

18. Khorana AA, Kuderer NM, Culakova E, et al. Development and validation of a predictive model for chemotherapy-associated thrombosis. Blood 2008;111(10):4902–7. doi: 10.1182/blood-2007-10-116327 [published Online First: 2008/01/25]

19. Kruger S, Haas M, Burkl C, et al. Incidence, outcome and risk stratification tools for venous thromboembolism in advanced pancreatic cancer - A retrospective cohort study. Thromb Res 2017;157:9–15. doi: 10.1016/j.thromres.2017.06.021

20. Faille D, Bourrienne MC, de Raucourt E, et al. Biomarkers for the risk of thrombosis in pancreatic adenocarcinoma are related to cancer process. Oncotarget 2018;9(41):26453–65. doi: 10.18632/oncotarget.25458 [published Online First: 2018/06/15]

21. Stahmeyer J, Stubenrauch S, Geyer S, et al. he frequency and timing of recurrent stroke—an analysis of routine health insurance data. Dtsch Ärztebl Int 2019;116:711–7.

22. Ryan AS, Ivey FM, Serra MC, et al. Sarcopenia and Physical Function in Middle-Aged and Older Stroke Survivors. Arch Phys Med Rehabil 2017;98(3):495–99. doi: 10.1016/j.apmr.2016.07.015 [published Online First: 2016/08/18]

23. Scherbakov N, von Haehling S, Anker SD, et al. Stroke induced Sarcopenia: muscle wasting and disability after stroke. Int J Cardiol 2013;170(2):89–94. doi: 10.1016/j.ijcard.2013.10.031 [published Online First: 2013/11/16]

24. Sanossian N, Djabiras C, Mack WJ, et al. Trends in cancer diagnoses among inpatients hospitalized with stroke. J Stroke Cerebrovasc Dis 2013;22(7):1146–50. doi: 10.1016/j.jstrokecerebrovasdis.2012.11.016 [published Online First: 2012/12/19]

25. Uemura J, Kimura K, Sibazaki K, et al. Acute stroke patients have occult malignancy more often than expected. Eur Neurol 2010;64(3):140–4. doi: 10.1159/000316764 [published Online First: 2010/07/30]

26. Cocho D, Gendre J, Boltes A, et al. Predictors of occult cancer in acute ischemic stroke patients. J Stroke Cerebrovasc Dis 2015;24(6):1324–8. doi: 10.1016/j.jstrokecerebrovasdis.2015.02.006 [published Online First: 2015/04/18]

27. Selvik HA, Thomassen L, Bjerkreim AT, et al. Cancer-Associated Stroke: The Bergen NORSTROKE Study. Cerebrovasc Dis Extra 2015;5(3):107–13. doi: 10.1159/000440730 [published Online First: 2015/12/10]

28. Weitbrecht WU, Kirchhoff D. [Long-term prognosis of cerebral infarct in comparison with a normal population]. Versicherungsmedizin 1995;47(2):46–9. [published Online First: 1995/04/01]

29. Meyer S, Verheyden G, Brinkmann N, et al. Functional and motor outcome 5 years after stroke is equivalent to outcome at 2 months: follow-up of the collaborative evaluation of rehabilitation in stroke across Europe. Stroke 2015;46(6):1613–9. doi: 10.1161/STROKEAHA.115.009421 [published Online First: 2015/05/09]

30. Drozdowska BA, Singh S, Quinn TJ. Thinking About the Future: A Review of Prognostic Scales Used in Acute Stroke. Front Neurol 2019;10:274. doi: 10.3389/fneur.2019.00274 [published Online First: 2019/04/06]

31. Pedersen A, Stanne TM, Redfors P, et al. Fibrinogen concentrations predict long-term cognitive outcome in young ischemic stroke patients. Res Pract Thromb Haemost 2018;2(2):339–46. doi: 10.1002/rth2.12078 [published Online First: 2018/07/27]

32. Swarowska M, Polczak A, Pera J, et al. Hyperfibrinogenemia predicts long-term risk of death after ischemic stroke. J Thromb Thrombolysis 2014;38(4):517–21. doi: 10.1007/s11239-014-1122-1 [published Online First: 2014/08/12]

33. Perlstein TS, Pande RL, Creager MA, et al. Serum total bilirubin level, prevalent stroke, and stroke outcomes: NHANES 1999-2004. Am J Med 2008;121(9):781–88 e1. doi: 10.1016/j.amjmed.2008.03.045 [published Online First: 2008/08/30]

34. Choi Y, Lee SJ, Spiller W, et al. Causal Associations Between Serum Bilirubin Levels and Decreased Stroke Risk: A Two-Sample Mendelian Randomization Study. Arterioscler Thromb Vasc Biol 2020;40(2):437–45. doi: 10.1161/ATVBAHA.119.313055 [published Online First: 2019/12/06]

35. Zhong P, Wu D, Ye X, et al. Association of circulating total bilirubin level with ischemic stroke: a systemic review and meta-analysis of observational evidence. Ann Transl Med 2019;7(14):335. doi: 10.21037/atm.2019.06.71 [published Online First: 2019/09/03]

36. Jorgensen ME, Torp-Pedersen C, Finer N, et al. Association between serum bilirubin and cardiovascular disease in an overweight high risk population from the SCOUT trial. Nutr Metab Cardiovasc Dis 2014;24(6):656–62. doi: 10.1016/j.numecd.2013.12.009 [published Online First: 2014/02/19]

37. Wang L, Li Y, Wang C, et al. C-reactive Protein, Infection, and Outcome After Acute Ischemic Stroke: A Registry and Systematic Review. Curr Neurovasc Res 2019;16(5):405–15. doi: 10.2174/1567202616666191026122011 [published Online First: 2019/11/19]

38. Martin AJ, Price CI. A Systematic Review and Meta-Analysis of Molecular Biomarkers Associated with Early Neurological Deterioration Following Acute Stroke. Cerebrovasc Dis 2018;46(5-6):230–41. doi: 10.1159/000495572 [published Online First: 2018/12/06]

39. Navi BB, Iadecola C. Ischemic stroke in cancer patients: A review of an underappreciated pathology. Ann Neurol 2018;83(5):873–83. doi: 10.1002/ana.25227 [published Online First: 2018/04/11]

40. Ellis D, Rangaraju S, Duncan A, et al. Coagulation markers and echocardiography predict atrial fibrillation, malignancy or recurrent stroke after cryptogenic stroke. Medicine (Baltimore) 2018;97(51):e13830. doi: 10.1097/MD.0000000000013830 [published Online First: 2018/12/24]

41. Nezu T, Kitano T, Kubo S, et al. Impact of D-dimer levels for short-term or long-term outcomes in cryptogenic stroke patients. J Neurol 2018;265(3):628–36. doi: 10.1007/s00415-018-8742-x [published Online First: 2018/01/27]

42. Chaudhary D, Abedi V, Li J, et al. Clinical Risk Score for Predicting Recurrence Following a Cerebral Ischemic Event. Front Neurol 2019;10:1106. doi: 10.3389/fneur.2019.01106 [published Online First: 2019/11/30]

43. Yanai H, Fraifeld VE. The role of cellular senescence in aging through the prism of Koch-like criteria. Ageing Res Rev 2018;41:18–33. doi: 10.1016/j.arr.2017.10.004 [published Online First: 2017/11/07]

44. Gonzalez-Meljem JM, Apps JR, Fraser HC, et al. Paracrine roles of cellular senescence in promoting tumourigenesis. Br J Cancer 2018;118(10):1283–88. doi: 10.1038/s41416-018-0066-1 [published Online First: 2018/04/20]

45. Xu M, Pirtskhalava T, Farr JN, et al. Senolytics improve physical function and increase lifespan in old age. Nat Med 2018;24(8):1246–56. doi: 10.1038/s41591-018-0092-9 [published Online First: 2018/07/11]

46. Baker DJ, Childs BG, Durik M, et al. Naturally occurring p16(Ink4a)-positive cells shorten healthy lifespan. Nature 2016;530(7589):184–9. doi: 10.1038/nature16932 [published Online First: 2016/02/04]

47. Baar MP, Brandt RMC, Putavet DA, et al. Targeted Apoptosis of Senescent Cells Restores Tissue Homeostasis in Response to Chemotoxicity and Aging. Cell 2017;169(1):132–47 e16. doi: 10.1016/j.cell.2017.02.031 [published Online First: 2017/03/25]

48. Justice JN, Nambiar AM, Tchkonia T, et al. Senolytics in idiopathic pulmonary fibrosis: Results from a first-in-human, open-label, pilot study. EBioMedicine 2019 doi: 10.1016/j.ebiom.2018.12.052

49. UNITY. UNITY Biotechnology Reports Promising Topline Data from Phase 1 First-in-human Study of UBX0101 in Patients with Osteoarthritis of the Knee, 2019.

50. Tanaka T, Biancotto A, Moaddel R, et al. Plasma proteomic signature of age in healthy humans. Aging Cell 2018;17(5):e12799. doi: 10.1111/acel.12799

51. Childs BG, Gluscevic M, Baker DJ, et al. Senescent cells: an emerging target for diseases of ageing. Nat Rev Drug Discov 2017;16(10):718–35. doi: 10.1038/nrd.2017.116 [published Online First: 2017/07/22]

52. Hisada Y, Mackman N. Cancer-associated pathways and biomarkers of venous thrombosis. Blood 2017;130(13):1499–506. doi: 10.1182/blood-2017-03-743211 [published Online First: 2017/08/16]

53. Moir JA, White SA, Mann J. Arrested development and the great escape--the role of cellular senescence in pancreatic cancer. Int J Biochem Cell Biol 2014;57:142–8. doi: 10.1016/j.biocel.2014.10.018 [published Online First: 2014/12/03]

54. Valenzuela CA, Quintanilla R, Moore-Carrasco R, et al. The Potential Role of Senescence As a Modulator of Platelets and Tumorigenesis. Front Oncol 2017;7:188. doi: 10.3389/fonc.2017.00188 [published Online First: 2017/09/13]

55. Posada-Duque RA, Barreto GE, Cardona-Gomez GP. Protection after stroke: cellular effectors of neurovascular unit integrity. Front Cell Neurosci 2014;8:231. doi: 10.3389/fncel.2014.00231 [published Online First: 2014/09/02]

56. Chan SL, Bishop N, Li Z, et al. Inhibition of PAI (Plasminogen Activator Inhibitor)-1 Improves Brain Collateral Perfusion and Injury After Acute Ischemic Stroke in Aged Hypertensive Rats. Stroke 2018;49(8):1969–76. doi: 10.1161/STROKEAHA.118.022056 [published Online First: 2018/07/12]

57. Garcia-Berrocoso T, Penalba A, Boada C, et al. From brain to blood: New biomarkers for ischemic stroke prognosis. J Proteomics 2013;94:138–48. doi: 10.1016/j.jprot.2013.09.005 [published Online First: 2013/09/26]

58. Mendioroz M, Fernandez-Cadenas I, Rosell A, et al. Osteopontin predicts long-term functional outcome among ischemic stroke patients. J Neurol 2011;258(3):486–93. doi: 10.1007/s00415-010-5785-z [published Online First: 2010/10/23]

59. Pan S, Chen R, Brand RE, et al. Multiplex targeted proteomic assay for biomarker detection in plasma: a pancreatic cancer biomarker case study. J Proteome Res 2012;11(3):1937–48. doi: 10.1021/pr201117w [published Online First: 2012/02/10]

60. Poruk KE, Firpo MA, Scaife CL, et al. Serum osteopontin and tissue inhibitor of metalloproteinase 1 as diagnostic and prognostic biomarkers for pancreatic adenocarcinoma. Pancreas 2013;42(2):193–7. doi: 10.1097/MPA.0b013e31825e354d [published Online First: 2013/02/15]

61. Alexander K, Yang HS, Hinds PW. Cellular senescence requires CDK5 repression of Rac1 activity. Mol Cell Biol 2004;24(7):2808–19. doi: 10.1128/mcb.24.7.2808-2819.2004 [published Online First: 2004/03/17]

62. Feldmann G, Mishra A, Hong SM, et al. Inhibiting the cyclin-dependent kinase CDK5 blocks pancreatic cancer formation and progression through the suppression of Ras-Ral signaling. Cancer Res 2010;70(11):4460–9. doi: 10.1158/0008-5472.CAN-09-1107 [published Online First: 2010/05/21]

63. Akinyemi R, Tiwari HK, Arnett DK, et al. APOL1, CDKN2A/CDKN2B, and HDAC9 polymorphisms and small vessel ischemic stroke. Acta Neurol Scand 2018;137(1):133–41. doi: 10.1111/ane.12847 [published Online First: 2017/10/05]

64. Cremin C, Howard S, Le L, et al. CDKN2A founder mutation in pancreatic ductal adenocarcinoma patients without cutaneous features of Familial Atypical Multiple Mole Melanoma (FAMMM) syndrome. Hered Cancer Clin Pract 2018;16:7. doi: 10.1186/s13053-018-0088-y [published Online First: 2018/03/16]

65. Wang T, Notta F, Navab R, et al. Senescent Carcinoma-Associated Fibroblasts Upregulate IL8 to Enhance Prometastatic Phenotypes. Mol Cancer Res 2017;15(1):3–14. doi: 10.1158/1541-7786.MCR-16-0192 [published Online First: 2016/09/30]

66. Chen J, Huang X, Halicka D, et al. Contribution of p16INK4a and p21CIP1 pathways to induction of premature senescence of human endothelial cells: permissive role of p53. Am J Physiol Heart Circ Physiol 2006;290(4):H1575–86. doi: 10.1152/ajpheart.00364.2005 [published Online First: 2005/10/26]

67. Tressera-Rimbau A, Arranz S, Eder M, et al. Dietary Polyphenols in the Prevention of Stroke. Oxidative medicine and cellular longevity 2017;2017:7467962. doi: 10.1155/2017/7467962

68. Angst E, Park JL, Moro A, et al. The flavonoid quercetin inhibits pancreatic cancer growth in vitro and in vivo. Pancreas 2013;42(2):223–9. doi: 10.1097/MPA.0b013e318264ccae

69. Yousefzadeh MJ, Zhu Y, McGowan SJ, et al. Fisetin is a senotherapeutic that extends health and lifespan. EBioMedicine 2018;36:18–28. doi: 10.1016/j.ebiom.2018.09.015

70. Khan FM, Zubek VB. Support Vector Regression for Censored Data (SVRc): A Novel Tool for Survival Analysis. Eighth IEEE International Conference on Data Mining. Pisa, Italy, 2008.

71. Ravichandran N, Suresh G, Ramesh B, et al. Fisetin, a novel flavonol attenuates benzo(a)pyrene-induced lung carcinogenesis in Swiss albino mice. Food and chemical toxicology : an international journal published for the British Industrial Biological Research Association 2011;49(5):1141–7. doi: 10.1016/j.fct.2011.02.005

72. Touil YS, Seguin J, Scherman D, et al. Improved antiangiogenic and antitumour activity of the combination of the natural flavonoid fisetin and cyclophosphamide in Lewis lung carcinoma-bearing mice. Cancer Chemother Pharmacol 2011;68(2):445–55. doi: 10.1007/s00280-010-1505-8

73. Khan N, Syed DN, Ahmad N, et al. Fisetin: a dietary antioxidant for health promotion. Antioxid Redox Signal 2013;19(2):151–62. doi: 10.1089/ars.2012.4901

74. Altman DG, McShane LM, Sauerbrei W, et al. Reporting Recommendations for Tumor Marker Prognostic Studies (REMARK): explanation and elaboration. PLoS Med 2012;9(5):e1001216. doi: 10.1371/journal.pmed.1001216 [published Online First: 2012/06/08]

75. Liu Y, Sanoff HK, Cho H, et al. Expression of p16(INK4a) in peripheral blood T-cells is a biomarker of human aging. Aging Cell 2009;8(4):439–48. doi: 10.1111/j.1474-9726.2009.00489.x

76. Ward-Caviness CK, Huffman JE, Everett K, et al. DNA methylation age is associated with an altered hemostatic profile in a multiethnic meta-analysis. Blood 2018;132(17):1842–50. doi: 10.1182/blood-2018-02-831347

77. Sousa-Santos AR, Amaral TF. Differences in handgrip strength protocols to identify sarcopenia and frailty - a systematic review. BMC Geriatr 2017;17(1):238. doi: 10.1186/s12877-017-0625-y [published Online First: 2017/10/19]

78. Oken MM, Creech RH, Tormey DC, et al. Toxicity and response criteria of the Eastern Cooperative Oncology Group. Am J Clin Oncol 1982;5(6):649–55. [published Online First: 1982/12/01]

79. van Swieten JC, Koudstaal PJ, Visser MC, et al. Interobserver agreement for the assessment of handicap in stroke patients. Stroke 1988;19(5):604–7. doi: 10.1161/01.str.19.5.604 [published Online First: 1988/05/01]

80. Rockwood K, Song X, MacKnight C, et al. A global clinical measure of fitness and frailty in elderly people. CMAJ 2005;173(5):489–95. doi: 10.1503/cmaj.050051 [published Online First: 2005/09/01]

81. Lyden P, Brott T, Tilley B, et al. Improved reliability of the NIH Stroke Scale using video training. NINDS TPA Stroke Study Group. Stroke 1994;25(11):2220–6. doi: 10.1161/01.str.25.11.2220 [published Online First: 1994/11/01]

82. Nasreddine ZS, Phillips NA, Bedirian V, et al. The Montreal Cognitive Assessment, MoCA: a brief screening tool for mild cognitive impairment. J Am Geriatr Soc 2005;53(4):695–9. doi: 10.1111/j.1532-5415.2005.53221.x [published Online First: 2005/04/09]

83. Herdman M, Gudex C, Lloyd A, et al. Development and preliminary testing of the new five-level version of EQ-5D (EQ-5D-5L). Qual Life Res 2011;20(10):1727–36. doi: 10.1007/s11136-011-9903-x [published Online First: 2011/04/12]

84. Snaith RP, Zigmond AS. The hospital anxiety and depression scale. Br Med J (Clin Res Ed) 1986;292(6516):344. doi: 10.1136/bmj.292.6516.344 [published Online First: 1986/02/01]

85. Ustun TB, Chatterji S, Kostanjsek N, et al. Developing the World Health Organization Disability Assessment Schedule 2.0. Bull World Health Organ 2010;88(11):815–23. doi: 10.2471/BLT.09.067231 [published Online First: 2010/11/16]

86. Lyons KD, Bakitas M, Hegel MT, et al. Reliability and validity of the Functional Assessment of Chronic Illness Therapy-Palliative care (FACIT-Pal) scale. J Pain Symptom Manage 2009;37(1):23–32. doi: 10.1016/j.jpainsymman.2007.12.015 [published Online First: 2008/05/28]

87. Sewtz C, Muscheites W, Kriesen U, et al. Questionnaires measuring quality of life and satisfaction of patients and their relatives in a palliative care setting-German translation of FAMCARE-2 and the palliative care subscale of FACIT-Pal. Ann Palliat Med 2018;7(4):420–26. doi: 10.21037/apm.2018.03.17 [published Online First: 2018/06/05]

88. Golicki D, Niewada M, Karlinska A, et al. Comparing responsiveness of the EQ-5D-5L, EQ-5D-3L and EQ VAS in stroke patients. Qual Life Res 2015;24(6):1555–63. doi: 10.1007/s11136-014-0873-7 [published Online First: 2014/11/27]

89. Ludwig K, Graf von der Schulenburg JM, Greiner W. German Value Set for the EQ-5D-5L. Pharmacoeconomics 2018;36(6):663–74. doi: 10.1007/s40273-018-0615-8 [published Online First: 2018/02/21]

90. Chuang LH, Cohen AT, Agnelli G, et al. Comparison of quality of life measurements: EQ-5D-5L versus disease/treatment-specific measures in pulmonary embolism and deep vein thrombosis. Qual Life Res 2019;28(5):1155–77. doi: 10.1007/s11136-018-2081-3 [published Online First: 2019/01/05]

91. Pickard AS, Neary MP, Cella D. Estimation of minimally important differences in EQ-5D utility and VAS scores in cancer. Health and quality of life outcomes 2007;5:70. doi: 10.1186/1477-7525-5-70

92. Hanley JA, McNeil BJ. The meaning and use of the area under a receiver operating characteristic (ROC) curve. Radiology 1982;143(1):29–36. doi: 10.1148/radiology.143.1.7063747 [published Online First: 1982/04/01]

93. Baur J, Moreno-Villanueva M, Kotter T, et al. MARK-AGE data management: Cleaning, exploration and visualization of data. Mech Ageing Dev 2015;151:38–44. doi: 10.1016/j.mad.2015.05.007 [published Online First: 2015/05/26]

94. Dereli O, Oguz C, Gonen M. Path2Surv: Pathway/gene set-based survival analysis using multiple kernel learning. Bioinformatics 2019;35(24):5137–45. doi: 10.1093/bioinformatics/btz446 [published Online First: 2019/05/31]

95. Buzdin A, Sorokin M, Garazha A, et al. Molecular pathway activation - New type of biomarkers for tumor morphology and personalized selection of target drugs. Semin Cancer Biol 2018;53:110–24. doi: 10.1016/j.semcancer.2018.06.003 [published Online First: 2018/06/24]

96. Warsow G, Greber B, Falk SS, et al. ExprEssence--revealing the essence of differential experimental data in the context of an interaction/regulation net-work. BMC Syst Biol 2010;4:164. doi: 10.1186/1752-0509-4-164 [published Online First: 2010/12/02]

97. Ernst M, Du Y, Warsow G, et al. FocusHeuristics - expression-data-driven network optimization and disease gene prediction. Sci Rep 2017;7:42638. doi: 10.1038/srep42638 [published Online First: 2017/02/17]

98. Hanzelmann S, Castelo R, Guinney J. GSVA: gene set variation analysis for microarray and RNA-seq data. BMC Bioinformatics 2013;14:7. doi: 10.1186/1471-2105-14-7 [published Online First: 2013/01/18]

99. Geistlinger L, Csaba G, Santarelli M, et al. Toward a gold standard for benchmarking gene set enrichment analysis. Brief Bioinform 2020 doi: 10.1093/bib/bbz158 [published Online First: 2020/02/07]

100. List M, Alcaraz N, Dissing-Hansen M, et al. KeyPathwayMinerWeb: online multi-omics network enrichment. Nucleic Acids Res 2016;44(W1):W98–W104. doi: 10.1093/nar/gkw373 [published Online First: 2016/05/07]

101. Neto E, Pratap A, Perumal T, et al. Using permutations to assess confounding in machine learning applications for digital health. ArXiv 2018; 1811.11920 or 1811.11920v1

102. Sorzano C, Tabas-Madrid D, Nunez F, et al. Sample Size for Pilot Studies and Precision Driven Experiments. ArXiv 2017; 1707.00222 or 1707.00222v2

103. Sauerbrei W, Schumacher M. A bootstrap resampling procedure for model building: application to the Cox regression model. Stat Med 1992;11(16):2093–109. doi: 10.1002/sim.4780111607 [published Online First: 1992/12/01]

104. Lin DY. Cox regression analysis of multivariate failure time data: the marginal approach. Stat Med 1994;13(21):2233–47. doi: 10.1002/sim.4780132105 [published Online First: 1994/11/15]

105. Binder H, Schumacher M. Allowing for mandatory covariates in boosting estimation of sparse high-dimensional survival models. BMC Bioinformatics 2008;9:14. doi: 10.1186/1471-2105-9-14 [published Online First: 2008/01/12]

106. Ishwaran H, Kogalur UB, Blackstone EH, et al. Random survival forests. Ann Appl Stat 2008;2(3):841–60. doi: 10.1214/08-AOAS169

107. Pi L, Halabi S. Combined Performance of Screening and Variable Selection Methods in Ultra-High Dimensional Data in Predicting Time-To-Event Outcomes. Diagn Progn Res 2018;2 doi: 10.1186/s41512-018-0043-4 [published Online First: 2018/11/06]

108. Ching T, Zhu X, Garmire LX. Cox-nnet: An artificial neural network method for prognosis prediction of high-throughput omics data. PLoS Comput Biol 2018;14(4):e1006076. doi: 10.1371/journal.pcbi.1006076 [published Online First: 2018/04/11]

109. Hao J, Kim Y, Kim TK, et al. PASNet: pathway-associated sparse deep neural network for prognosis prediction from high-throughput data. BMC Bioinformatics 2018;19(1):510. doi: 10.1186/s12859-018-2500-z

110. Yousefi S, Amrollahi F, Amgad M, et al. Predicting clinical outcomes from large scale cancer genomic profiles with deep survival models. Sci Rep 2017;7(1):11707. doi: 10.1038/s41598-017-11817-6111.

111. Bass A, Storey J. bioRxiv 2019 doi: 10.1101/571992

112. Moeller S, Saul N, Cohen AA, et al. Healthspan pathway maps in C. elegans and humans highlight transcription, prolifera-tion/biosynthesis and lipids. bioRxiv 2018

113. Motwani HV, Frostne C, Tornqvist M. Parallelogram based approach for in vivo dose estimation of genotoxic metabolites in humans with relevance to reduction of animal experiments. Sci Rep 2017;7(1):17560. doi: 10.1038/s41598-017-17692-5

114. Kienhuis AS, van de Poll MC, Wortelboer H, et al. Parallelogram approach using rat-human in vitro and rat in vivo toxicogenomics predicts acetaminophen-induced hepatotoxicity in humans. Toxicol Sci 2009;107(2):544–52. doi: 10.1093/toxsci/kfn237

115. Taroni JN, Grayson PC, Hu Q, et al. MultiPLIER: A Transfer Learning Framework for Transcriptomics Reveals Systemic Features of Rare Disease. Cell Syst 2019;8(5):380–94 e4. doi: 10.1016/j.cels.2019.04.003 [published Online First: 2019/05/24]

116. Schussler-Fiorenza Rose SM, Contrepois K, Moneghetti KJ, et al. A longitudinal big data approach for precision health. Nat Med 2019;25(5):792–804. doi: 10.1038/s41591-019-0414-6 [published Online First: 2019/05/10]

117. Avelar RA, Ortega JG, Tacutu R, et al. A Multidimensional Systems Biology Analysis of Cellular Senescence in Ageing and Disease. bioRxiv 2019

118. Demaria M, O’Leary MN, Chang J, et al. Cellular Senescence Promotes Adverse Effects of Chemotherapy and Cancer Relapse. Cancer Discov 2017;7(2):165–76. doi: 10.1158/2159-8290.CD-16-0241 [published Online First: 2016/12/17]

119. Fulop T, Larbi A, Dupuis G, et al. Immunosenescence and Inflamm-Aging As Two Sides of the Same Coin: Friends or Foes? Front Immunol 2017;8:1960. doi: 10.3389/fimmu.2017.01960

